# The vanishing white matter registry: a case study of historical controls in clinical trial design

**DOI:** 10.64898/2026.08.19.26360800

**Authors:** Romy J. van Voorst, Elia Gonzato, Eline M.C Hamilton, Menno D. Stellingwerf, Merel C. Postema, Johannes Berkhof, Rik van Eekelen, Marjo S. van der Knaap

## Abstract

**Background:** Therapy development in ultra-rare, progressive and fatal diseases like vanishing white matter (VWM) is hampered by very low patient numbers and ethical constraints regarding placebo-controlled studies. Under such conditions, standard randomized controlled trials may not be feasible. The use of historical control information could be part of a solution, but would require extra considerations regarding selection of patients and choice of endpoints. We used the VWM registry as a case study to outline key methodological considerations for informing trial design in ultra-rare disease.

**Methods:** The study included 462 patients, available in the VWM registry. Prospective clinical data were collected since 2004 using VWM-specific questionnaire and Health Utility Index (HUI) assessments, while retrospective data from clinical charts were available from 1988 on. We evaluated methodological aspects relevant to trial design, including patient selection, drift in the disease course over time, endpoint selection, and clinically relevant stratification into subgroups.

**Results:** Regarding patient selection, patients with comorbidities impacting disease course, and pre-symptomatic individuals without clinical onset were considered not suitable as historical controls. After excluding patients before 1991, we found no evidence of drift in the disease course from 1991 onwards. Regarding choice of endpoints, episodes of rapid decline were relatively infrequent and occurred mostly at disease onset, limiting their usefulness as trial endpoint. Multi-state modelling and clinical evaluation showed ambulation as preferable endpoint over survival. For longitudinal HUI multiscores, baseline imputation allowed modelling of early disease. The scores showed a distinct ordering, reflecting the association between multi-domain function and disease progression. Regarding stratification, the combination of data-driven analyses and clinical expertise informed revised age of onset groups. Females showed later onset and milder disease; adjustment for age of onset eliminated the effect of sex.

**Conclusion:** This case study provides key considerations for evaluating registry data as historical control and demonstrates how these considerations can inform clinical trial design in ultra-rare diseases.

## Introduction

Vanishing White Matter (VWM, OMIM #603896) is a leukodystrophy characterized by degeneration of CNS white matter. It is ultra-rare with estimated live birth incidence of approximately 1:100,000 and a prevalence of approximately 1.3: 1,000,000.^1,2^ VWM is caused by biallelic pathogenic variants in the EIF2B1–5 genes, which encode eukaryotic initiation factor 2B (eIF2B), a key factor in mRNA translation and regulation of the integrated stress response (ISR)^3^. With the ISR defect, patients with VWM are susceptible to physical stressors, which may provoke episodes of rapid decline.^2–4^ VWM may present at any age, but two-third of the patients has an onset below 6 years of age.^1,2^ The disease course varies widely between patients, with age of onset being the most reliable predictor of progression, i.e. earlier onset is associated with a faster progression and earlier death.^2^

In the context of ultra-rare diseases like VWM, where recruiting sufficiently large sample sizes for randomized controlled trials with desirable operating characteristics (e.g. adequate type 1 error control and statistical power) is unfeasible, and the rapidly progressive and fatal disease course in young children make placebo controls ethically problematic^5^, alternative designs must be considered. In such cases, availability of longitudinal follow-up information of as many untreated patients as possible, documenting the natural history course, is essential.^41,6,7^ In randomized controlled trials with at least two treatment arms, historical control data can augment the total sample size and achieve the same statistical power while enrolling a lower number of patients.^1,8^ In single-arm trials, historical control data may enable assessment of comparative effectiveness.^7,9,10^

The VWM patient registry could serve as a source of historical data to inform clinical trial design. Data collection started prospectively in 2004^4^, but also retrospective data were collected from patients seen since 1988^2^, covering almost 40 years. We used the VWM registry as a methodological case study to evaluate the feasibility and limitations of historical controls in clinical trials for ultra-rare diseases, addressing several crucial points. The current study adds to a previous study on the natural history of VWM^2^, by including a larger number of patients and longer follow-up, enabling examination of (1) special subpopulations and their suitability as historical controls, (2) potential drift in disease course limiting comparability between past and contemporary cohorts, (3) suitable end points, and (4) necessary stratification of patients.

## Patient and Methods

### Study Design

This longitudinal observational study included data from the VWM registry. To be enrolled in the registry, patients must have an MRI- and DNA-confirmed diagnosis.^11^ Written informed consent was obtained from participating patients or guardians. The study was approved by the ethics committee of VU University Medical Center, Amsterdam in 2003 (2003.194) and 2018 (2018.588). The HMA-EMA Data Quality Framework^12^ was used to assess data quality. Data were collected via Castor EDC/CDMS with automated checks for inconsistencies and expert review of major changes in key items.^5^

The VWM registry prospectively collects data from January 2004 onwards. The protocol for data collection includes administering online questionnaires and inventories four times per year for patients under 4 years of current age, two times per year for patients that are 4 to under 8 years, and once per year for patients 8 years and older. The registry also contains retrospectively collected data from 1988 and retrospective data from patients who entered the registry at a later disease stage. In this study, we used data collected until the 1^st^ of July 2025, referred to as the date of study closure. When patients enrolled in a clinical trial, they were excluded from participation in the registry from that moment on.

### Customized VWM-specific questionnaires

We obtained information from a customized VWM-specific clinical questionnaire.^2^ This questionnaire was completed by the patient, family member, physician or any combination thereof. We included the following clinical characteristics: date of birth, country of residence, sex, age of onset, age at follow-up, disease duration at follow-up, time of follow-up at questionnaire, and number of questionnaires.

Age of onset was defined as the time point at which clinical examination revealed the first neurological sign attributable to the disease (most often signs of ataxia or spasticity), as confirmed by physicians’ medical documentation. White matter abnormalities on MRI per se were not considered indicative of clinical onset.^7^ Isolated ovarian insufficiency, developmental delay, mental retardation, cataract and transient neurological signs, such as seizures, migraine attacks, vertigo, and syncope, were not classified as neurological disease onset.

Disease duration was defined as the interval between age of onset and age at last recorded follow-up. Age of onset was used as the starting point, i.e. time zero, because it has the clearest definition that reflects the natural disease course of VWM. In VWM and other leukodystrophies, the specific DNA-confirmed diagnosis used to be lacking or delayed for many years up to decades^13^, has become faster in more recent years^14,15^, and is currently increasingly established in the presymptomatic stage, making that the date of diagnosis is not a uniform clinical event in the disease course for each patient. Likewise, registry enrollment happens at a variable point in the disease course that in more recent years tends to become earlier in the disease course.^1,7^ Date of diagnosis and date of registry enrolment were therefore not considered suitable as starting point.

Disease progression was assessed using the clinical endpoints ambulation and overall survival. Patients were classified as having lost ambulation without support when they no longer could walk without assistive devices or another person’s help. Loss of ambulation with support was defined as full wheelchair dependence. Patients who never achieved ambulation without or with support were excluded from ambulation analyses. The occurrence of episodes of acute decline were documented, as reported by families and physicians.

### The Health Utilities Index

The Health Utilities Index (HUI) is a validated inventory assessing health-related quality of life and was previously used in a VWM natural history study^2^. For the registry, the HUI Mark 3 (HUI3) proxy version was administered from age 2 years onward. It comprises eight attributes (Vision, Hearing, Speech, Ambulation, Dexterity, Emotion, Cognition, and Pain), rated on 5- or 6-level scales. HUI3 single-attribute scores ranging from 0 (severe impairment) to 1 (no impairment) were used to calculate a multiscore over all eight attributes, ranging from −0.36 (with a score below 0 indicating worse than death in the opinion of the general population) to 1.00 (perfect health).^16^ Similar to the VWM natural history study^2^, we assigned a score of −0.50 instead of 0 to death to account for the positive emotional well-being observed in some patients who had negative utility scores during life.

### Statistical analysis

#### General definitions

The time-axis for longitudinal analyses was the disease duration. Demographic data were described using summary statistics: mean and standard deviation (SD) for continuous variables that were normally distributed, or median and min-max range otherwise. Categorical variables were reported as counts and percentages (n [%]). Identified non-linear relationships were modeled using polynomial terms or natural or restricted cubic splines, after which the best-fitting models were selected. Model fit was evaluated using the lowest Bayesian Information Criterion (BIC) and Akaike Information Criterion (AIC).^17,18^ We identified non-proportional hazards using scaled Schoenfeld residuals.^19^

#### Drift

To analyze drift in disease progression regarding loss of ambulation and survival, we used Cox proportional hazards regression with year of onset and age of onset as covariates. Model performance was validated using Kaplan–Meier curves, for which we grouped year of onset per five years to increase patient numbers per group. Following analysis of the full cohort, a second model was restricted to patients with disease onset from 2002 on, when genetic testing and standardized disease management guidelines were available.^20, 21^The second model was used for all subsequent analyses.

#### Disease stage transitions

Endpoints must be clinically meaningful and statistically robust. We assessed the endpoints of ambulation with support and survival. From a statistical perspective, a multi-state model was applied to characterize disease transitions over time for the states ‘independent ambulation’, ‘having lost ambulation with support’, and ‘death’. Given its reported strong predictive value for disease progression^2^, age of onset was included as covariate to assess its effect on transitions between stages.

#### Age of onset categories

We reassessed the categories for age of onset as previously defined in the natural history study with more data.^2^ We first applied recursive partitioning on the covariate age of onset to identify data-driven categorization for the endpoint ambulation with support. We used the function rpart picks nodes based on the log-rank statistics. To check robustness and consistency of this data-driven procedure, we bootstrapped the procedure 1000 times to determine how often categories were picked by the approach. Bootstrapped results were used for visualization only. We considered the most frequently picked categories to be the data-driven categories. Final agreement on the new age of onset categories was reached combining these data-driven results with clinical expertise (MSvdK).

#### Sex

Differences in age of onset between sexes were assessed using the Wilcoxon rank-sum test. A Cox proportional-hazards model adjusted for age of onset was used to evaluate the effect of sex on ambulation with support and overall survival, adjusted for age of onset using restricted cubic splines with three knots.

#### Episodes of rapid decline

Regarding the frequency of reported episodes of rapid decline, the mode, median and range of number of episodes per patient were calculated. We assessed the proportion of patients presenting with an episode of rapid decline at disease onset and associated the age of onset to this proportion.

#### HUI multiscores

We summarized HUI multiscores by ambulation states (independent walking, walking with support, no walking, and death) using the mean, standard deviation, median, and first and third quartiles. Spaghetti plots were created to visualize the repeated measurements of HUI multiscores over time. The HUI plots were stratified by the new categories of age of onset identified in the previous analysis. We then modelled the trajectory in time of the HUI multiscores using the different functional forms (described in the General definitions paragraph), with natural cubic splines. We used the best-performing model to estimate the average HUI multiscore over time for each category of age of onset.

As the HUI multiscore was rarely assessed right before disease onset and our time axis starts at age of onset, there were many missing data on what could be considered the baseline measurement. We analyzed the effect of imputation of baseline scores at disease onset corresponding to age-matched reference scores from HUI3 reference population data.^22,23^ For patients below 5 years, we applied the reference score of 0.92 for the lowest age, i.e. 5 years. As sensitivity analysis, we repeated analyses without imputing these baseline measurements: first visualizing in spaghetti plots, then using the best-performing model to estimate HUI multiscores over the years between 2002 and 2025.

Missing data later during follow-up were addressed using inspection and logical deduction per published guidelines.^24^ Inventories with more than two missing attributes were excluded (10% of cases). For up to two missing values, we applied hot deck imputation based on pattern similarity. Hot deck imputations were calculated for the HUI3 attribute scores on Cognition in 0.7% of inventories, 0.5% on Emotion, 0.4% on Hearing and 0.1%, on Vision, Ambulation, Dexterity and Speech.

#### Software

Data analysis and visualization were conducted using R version 4.4.3 (R Core Team (2025); R Foundation for Statistical Computing, Vienna, Austria).

## Results

### Patients and subpopulations in the VWM registry

On July 1, 2025, 462 patients were available for the study. Four special subpopulations, for which eligibility as historical control could be questioned, were identified within the registry.

The first identified subpopulation had important comorbidities. Ten patients had biliary atresia, Down syndrome, glutaric aciduria type 1, encephalocele and cortical dysplasia, galactosemia, Leigh syndrome due to biallelic LRPPRC mutations (3 siblings), Sjögren-Larsson syndrome, and perinatal asphyxia. They were excluded from further analyses because the probable effect of the comorbidity on the disease course.

The second subpopulation comprised 13 patients without clinical neurological signs attributable to VWM. Four had not had any disease signs: two patients, 5 and 3 years at last follow-up, had genetic testing because of an affected sibling; two patients, 6 and 21 years at last follow-up, were identified by MRI because of head trauma or headache. Four patients had been identified by MRI for transient neurological signs: vertigo, syncope, unprovoked dizziness, and one-sided vision loss. They were 13, 24, 12, and 50 years, respectively, at last follow-up. The other five patients had been identified because of isolated symptoms: cataract in two siblings, aged 29 and 30 years at last follow-up, two individuals because of learning and behavioral difficulties aged 11 and 10, and dysmorphic features in one person aged 11 years. These patients were excluded from further analyses because they had no disease onset.

The third subpopulation were patients with transient neurological signs attributable to VWM and full clinical recovery. Seven patients experienced an episode of neurological signs following a stress-provoked trigger, recovered after this episode and remained asymptomatic so far. In three of these patients the episodes were triggered by a head trauma at ages 4, 8, and 2 years. At last follow-up they were 21, 13, 10 years old, respectively. Three patients presented with an episode triggered by a febrile infection at ages 4, 5, and 2 years. At last follow-up, these patients were 14, 14, and 10 years old, respectively. One patient experienced an episode triggered by sleep deprivation at the age of 18 and was 41 years at last follow-up. During these episodes, patients experienced a range of neurological signs and symptoms, including combinations of seizures, headaches, vomiting, vision loss, drowsiness and lowered consciousness, together with abnormal findings at neurological examination. All signs and symptoms resolved after the episodes. These patients were excluded from analysis, because they were asymptomatic after the transient presentation.

The fourth subpopulation comprised 16 patients that presented with an episode of acute neurological decline ending in death. Most patients (88%, 14/16) had early-infantile disease onset, which is associated with rapid disease progression^2^. Notably, two patients, ages 16 and 17 years, who were asymptomatic, experienced an episode of rapid decline resulting in death. The trigger in both was a respiratory tract infection. These 16 patients were included in further analyses.

After the exclusions, a total of 432 patients (204 males and 228 females) remained in the study.

### Clinical and Demographic Characteristics

Genetic analysis revealed that seven patients had biallelic variants in EIF2B1, 69 in EIF2B2, 36 in EIF2B3, 37 in EIF2B4, and 283 in EIF2B5. Median age at last follow-up was 13 years (range 3 months–78 years; n=427), median disease duration 8 years (range 1 week–48 years; n=423), and median age at onset 3 years (range 0–74 years; n=427). Patients resided in Africa (n=1), Australia and New Zealand (n=14), Asia (n=32), South America (n=46), North America (n=78) and Europe (n=261). Patients with missing data were selectively included based on available information. The number of patients included in each analysis is reported in parentheses or in the corresponding tables.

### Available data

Questionnaires and inventories are distributed according to the protocol schedule. Although not all participants completed every assessment at each time point, a substantial longitudinal dataset was obtained (Figure 1). VWM-specific questionnaires were most frequently available (n=987), followed by HUI inventories (n=933). Time to follow-up between questionnaires and inventories ranged from 0 to 21 years (median 4 months).

**Figure 1.**
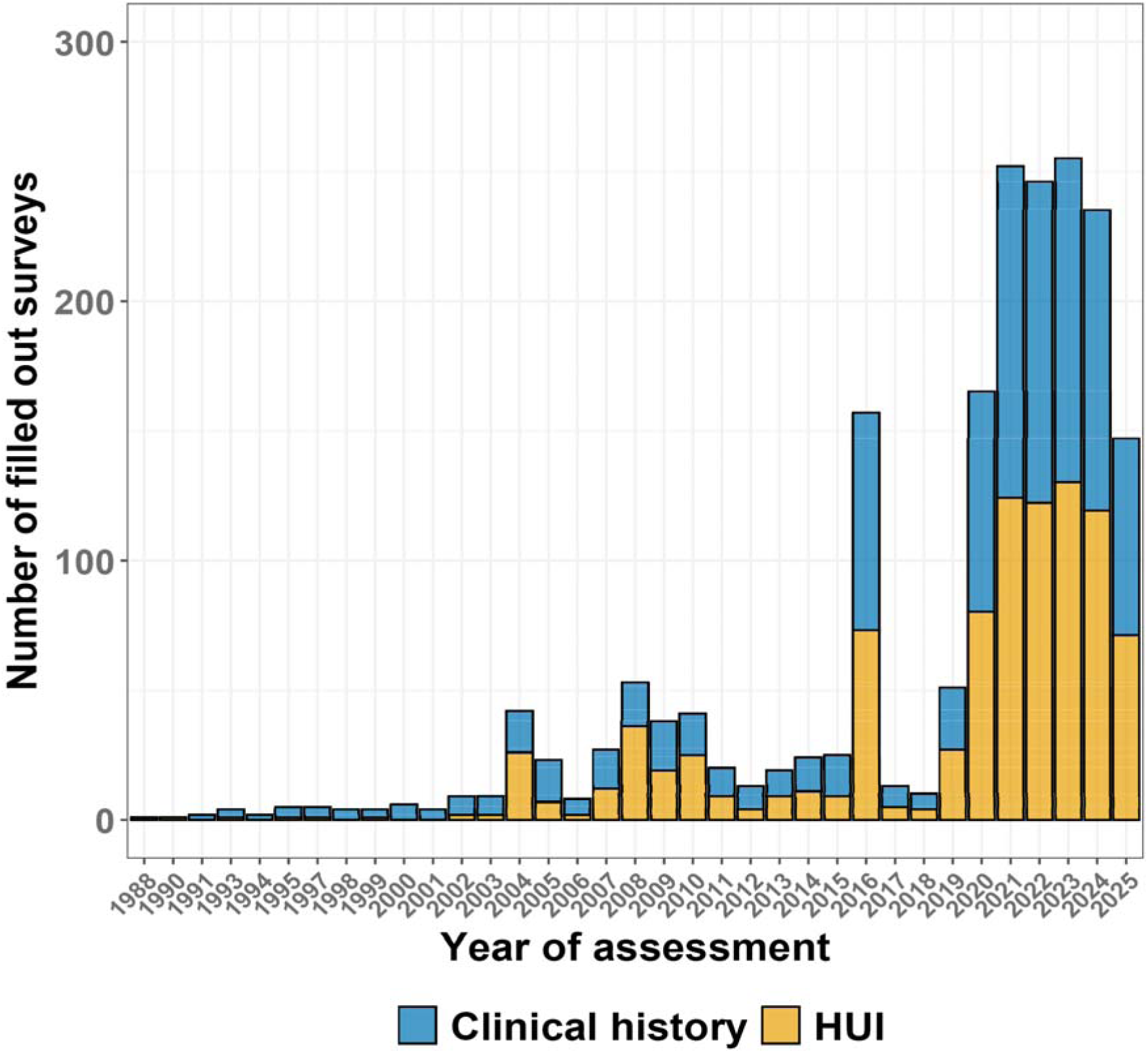
Frequency of the different customized VWM-specific questionnaire and HUI inventory

### Drift

Ambulation and survival are preferred outcome measures in leukodystrophy therapy trials^5,25^. They were assessed for drift. Information on ambulation was available on 404 patients and missing in 28. Unsupported ambulation was achieved in 341 of 404 patients, while 63 patients never walked without support. In total, 165 out of 432 patients had passed away at last follow-up. The Cox model indicated a trend toward milder disease course regarding loss of ambulation without or with support and death among the oldest historical controls (Table 1). Kaplan–Meier analyses revealed a markedly different disease course in patients with onset as early as 1975 compared with patients from 1991 onwards (Figure 2). After excluding historical controls before 1991, the Cox model provided no evidence for drift thereafter (Table 1, Figure 2 and Supplementary Figure 1), although this test does not exclude presence of drift altogether.

**Figure 2.**
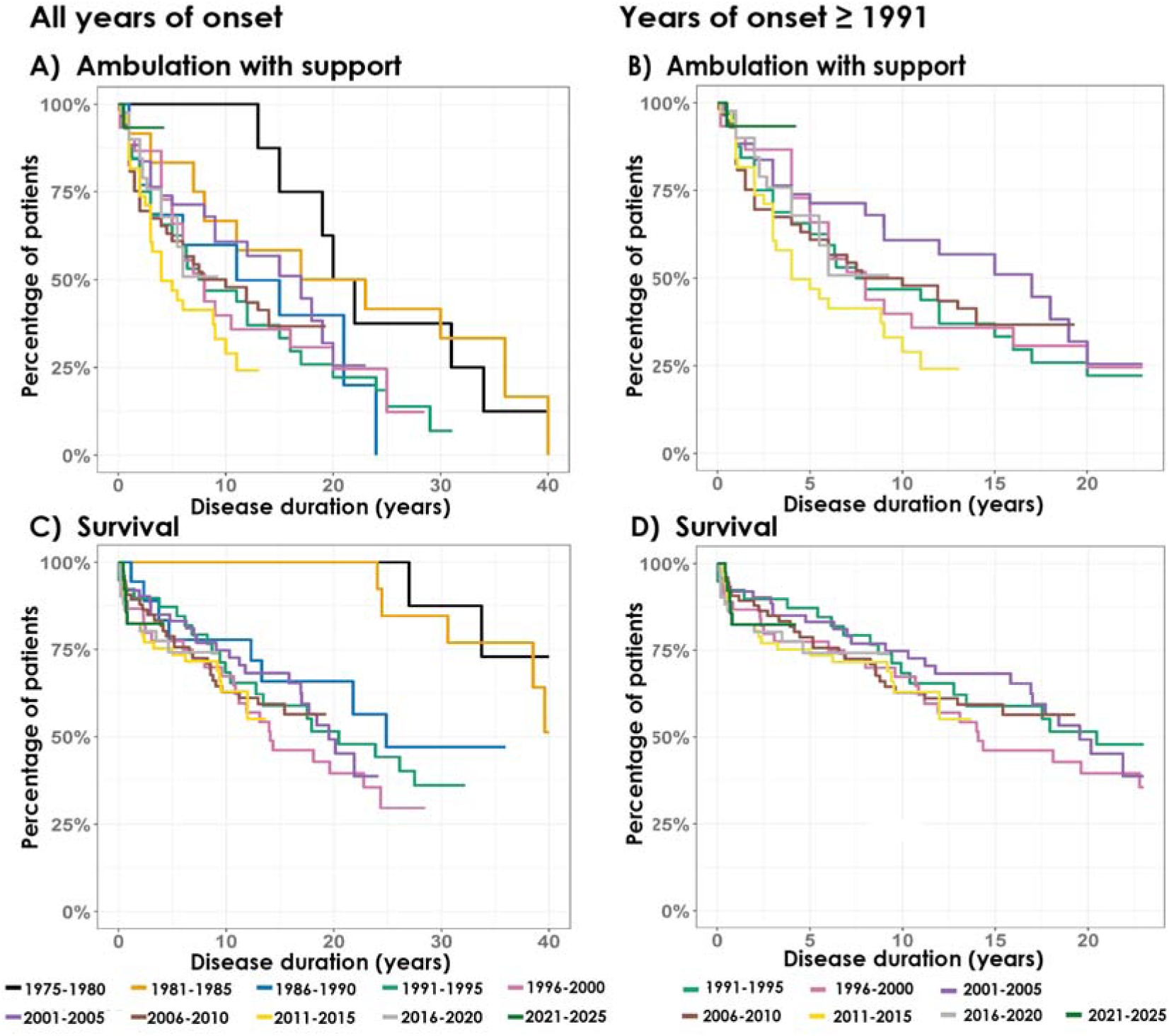
Kaplan–Meier curves with year of onset and loss of ambulation with support and overall survival A) Overall time to loss of ambulation with support by year of onset: 1975-1980 (n=8); 1981-1985 (n=12), 1986–1990 (n=13), 1991–1995 (n=32), 1996–2000 (n=30), 2001-2005 (n=44), 2006–2010 (n=58), 2011–2015 (n=39), 2016–2020 (n=40), 2021–2025 (n=36); B) Time to loss of ambulation with support in patients with onset from 1991 onwards: 1991-1995 (n=32), 1996-2000 (n=30), 2001-2005 (n=44), 2006–2010 (n=58), 2011–2015 (n=39), 2016–2020 (n=40), 2020–2025 (n=36); C) Overall survival by year of onset: 1975-1980 (n=8); 1981-1985 (n=14), 1986–1990 (n=18), 1991–1995 (n=39), 1996–2000 (n=45), 2001-2005 (n=63), 2006–2010 (n=75), 2011–2015 (n=58), 2016–2020 (n=51), 2021–2025 (n=45); D) Overall survival in patients with onset from 1991 onwards: 1991-1995 (n=39), 1996-2000 (n=45), 2001-2005 (n=63), 2006–2010 (n=75), 2011–2015 (n=58), 2016–2020 (n=51), 2020–2025 (n=45).

**Table 1.**
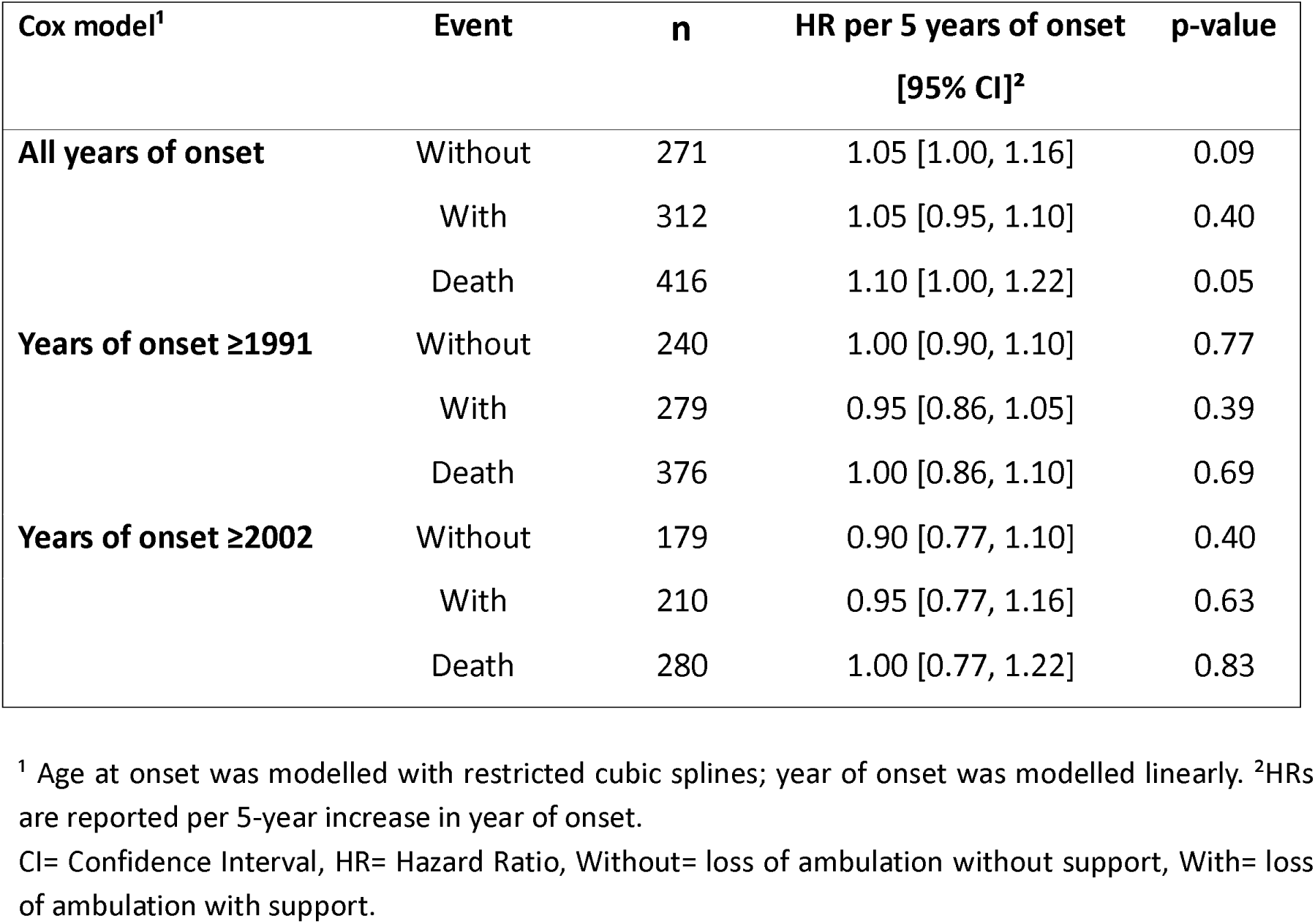
Effect of onset year on ambulation loss and overall survival, adjusted for age at onset.

| Cox model<sup>1</sup> | Event | n | HR per 5 years of onset<br>[95% CI]<sup>2</sup> | p-value |
| --- | --- | --- | --- | --- |
| **All years of onset** | Without | 271 | 1.05 [1.00, 1.16] | 0.09 |
|  | With | 312 | 1.05 [0.95, 1.10] | 0.40 |
|  | Death | 416 | 1.10 [1.00, 1.22] | 0.05 |
| **Years of onset ≥1991** | Without | 240 | 1.00 [0.90, 1.10] | 0.77 |
|  | With | 279 | 0.95 [0.86, 1.05] | 0.39 |
|  | Death | 376 | 1.00 [0.86, 1.10] | 0.69 |
| **Years of onset ≥2002** | Without | 179 | 0.90 [0.77, 1.10] | 0.40 |
|  | With | 210 | 0.95 [0.77, 1.16] | 0.63 |
|  | Death | 280 | 1.00 [0.77, 1.22] | 0.83 |

<sup>1</sup> Age at onset was modelled with restricted cubic splines; year of onset was modelled linearly. <sup>2</sup>HRs are reported per 5-year increase in year of onset.
CI= Confidence Interval, HR= Hazard Ratio, Without= loss of ambulation without support, With= loss of ambulation with support.

Alternatively, statistical analyses can be combined with knowledge of the disease and changes over time for selection of patient groups. The genes that are associated with VWM were published in 2001 and 2002, making the diagnosis of VWM more straightforward and standardized than before^21,26^. In addition, as the relationship between the associated genes and the cell stress response was unveiled, the usual care for patients changed, introducing protocols aimed at avoiding physical stressors^11^. This could be a reasonable combined argument to choose controls from the registry from 2002 onwards. After excluding historical controls before 2002, the Cox model provided no evidence for drift thereafter (Table 1, Figure 2).

### Disease stage transitions

Age of onset significantly influenced the transition from independent ambulation to loss of ambulation with support, with younger age of onset patients transitioning faster than later onset patients (HR=0.97 per increasing year, 95%CI [0.95, 0.99], n=169, p=0.0007). Age of onset did not have a significant effect on the transition from ambulation with support to death (HR=1.02 per increasing year, 95%CI [0.99, 1.04], n=77, p = 0.17). Predictions of the disease transitions are available (Supplementary Figure 2).

### Age of onset categories

Evaluation of the previously defined^2^ age of onset categories confirmed overlap in disease progression for ambulation with support and overall survival between the categories (Figure 3). Recursive partitioning revealed three distinct age of onset categories for ambulation with support: 1-<2 years, 2-<3 years and ≥3 years. Based on clinical relevance, the ≥3 years group was further subdivided into 3–<7 years and ≥7 years. In analysis of survival, we also included severely affected patients who never achieved ambulation with support, resulting in the following age of onset groups: <1 year, 1–<2 years, 2–<3 years, 3–<7 years, and ≥7 years. Among all onset groups, the patients with an age of onset between 3-<7 years constituted the largest group for both ambulation with support (35%; n=73/210) and overall survival (29%; n=82/280).

**Figure 3.**
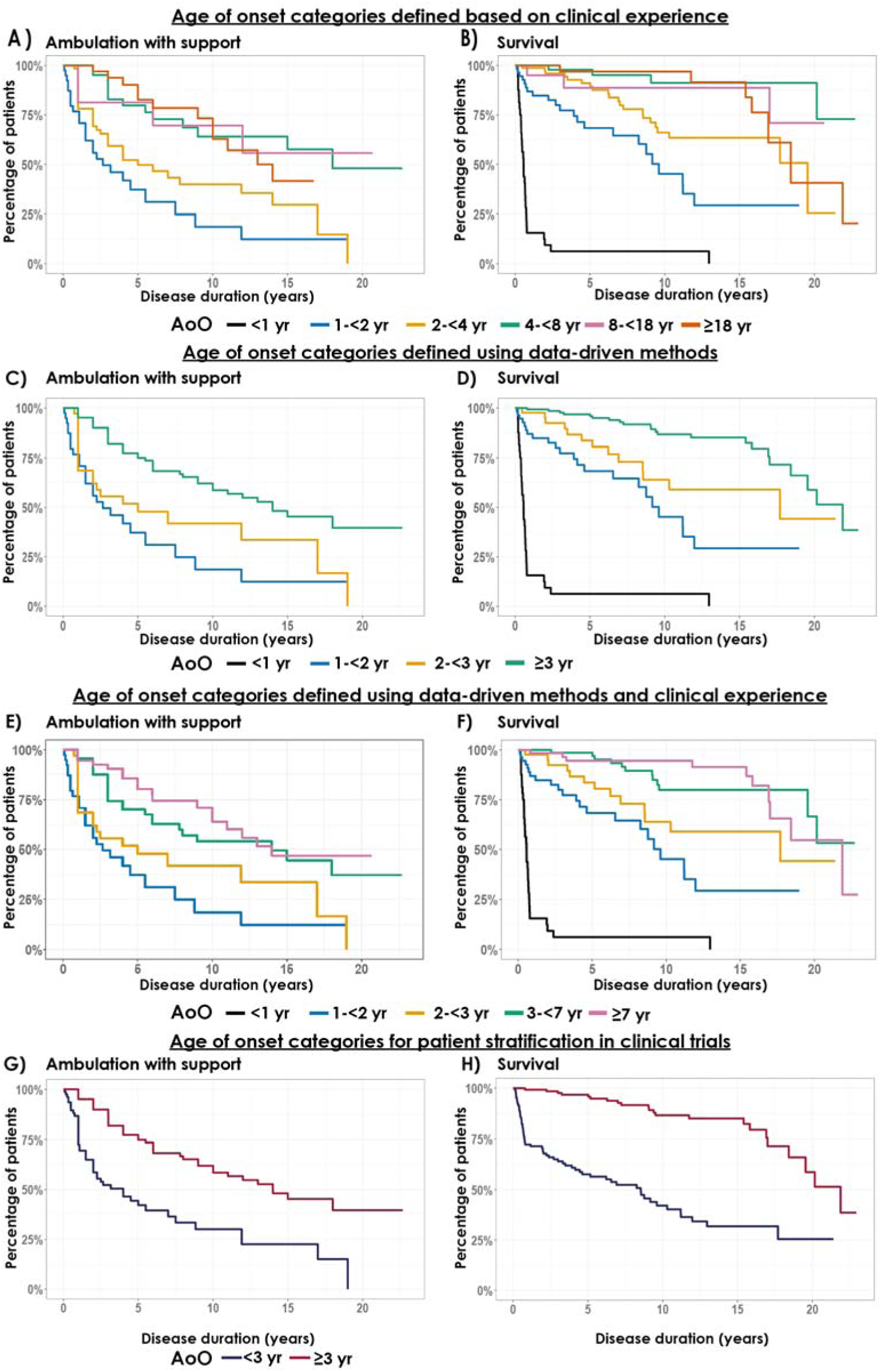
Kaplan-Meier curves with age of onset and loss of ambulation with support and overall survival Clinical defined age of onset categories for A)) ambulation with support 1-<2 years (n=39), 2-<4 years (n=72), 4-<8 years (n=47), 8-<18 years(n=18), ≥18 years (n=34) and B) overall survival <1 years (n=34), 1-<2 years (n=55), 2-<4 years (n=83), 4-<8 years (n=51), 8-<18 years(n=22), ≥18 years (n=35). Data driven age of onset categories for C) ambulation with support: 1--<2 years (n=39), 2-<3 years (n=40) ≥3 years (n=131) and D) survival <1 years (n=34), 1--<2 years (n=55), 2-<3 years (n=45) ≥3 years (n=146). Age of onset categories combining data driven method and clinical experience for E) ambulation with support ( 1–<2 years (n=39), 2–<3 years (n=40), 3–<7 years (n=73), and ≥7 years (n=58)) and F) survival (<1 year (n=34), 1–<2 years (n=55), 2–<3 years (n=45), 3–<7 years (n=82), and ≥7 years (n=64)). Age of onset categories for patient stratification in clinical trials G) ambulation with support <3 years (n=79) and ≥3 years (n=131) and H) survival <3 years (n=134) and ≥3 years (n=146). AoO= age of onset in years.

Kaplan–Meier curves indicated that a small number of patients, particularly one patient in the <1 year onset group, had a much longer survival than the other patients in the same cohort.

### Sex

Females had a higher average age of onset in years (mean= 9.6, SD ± 14.3; n=145) than males (mean = 5.2, SD ± 9.0; n=141). The median age of onset of females was 3.0 (range= 0-74) and 2.0 for males (range= 0-50). The Wilcoxon rank sum test for the difference in age of onset between females and males was significant, providing evidence that males tend to have an earlier onset than females (p=0.003). Kaplan Meier curves showed a difference between the sexes; however the effect of sex on ambulation with support did not reach statistical significance. In contrast, the effect of sex on overall survival was statistically significant (HR 1.81, 95% [1.19, 2.27], n==88/283, p=0.006) (Figure 4).

**Figure 4.**
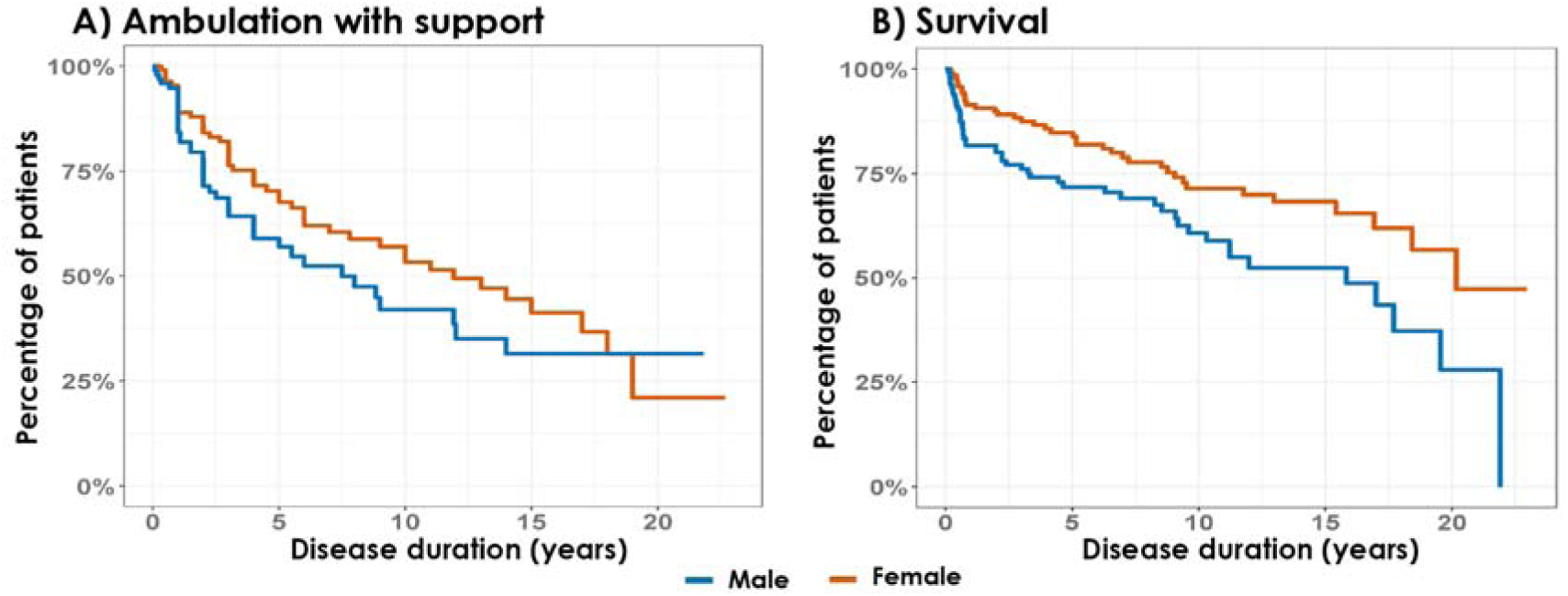
Kaplan-Meier curves for sex and loss of ambulation with support and death The curves show the effect of sex on A) loss of ambulation with support (n=112 females; n= 98 males) and B) overall survival (n=143 females; n= 137 males).

After correcting for age of onset, the remaining effect of sex on ambulation with support (HR=1.47, 95%CI= [0.96, 2.25], n=98/210, p=0.076) and survival (HR=1.47, 95%CI=[0.95, 2.27], n=137/280, p=0.081) was no longer statistically significant.

### Episodes of rapid decline

Overall, 85% of patients (331/388) experienced at least one episode of acute decline during the disease course. In these patients, the median number of episodes per patient was 2 (n=247, range: 1– 39) and the mode was 1. Approximately 49% of patients (122/247) experienced an episode at disease onset, most commonly in patients with an age of onset <7 years (95/122) (Figure 5). Among the patients with an episode at disease onset, 15% (37/247) had no subsequent episodes.

**Figure 5.**
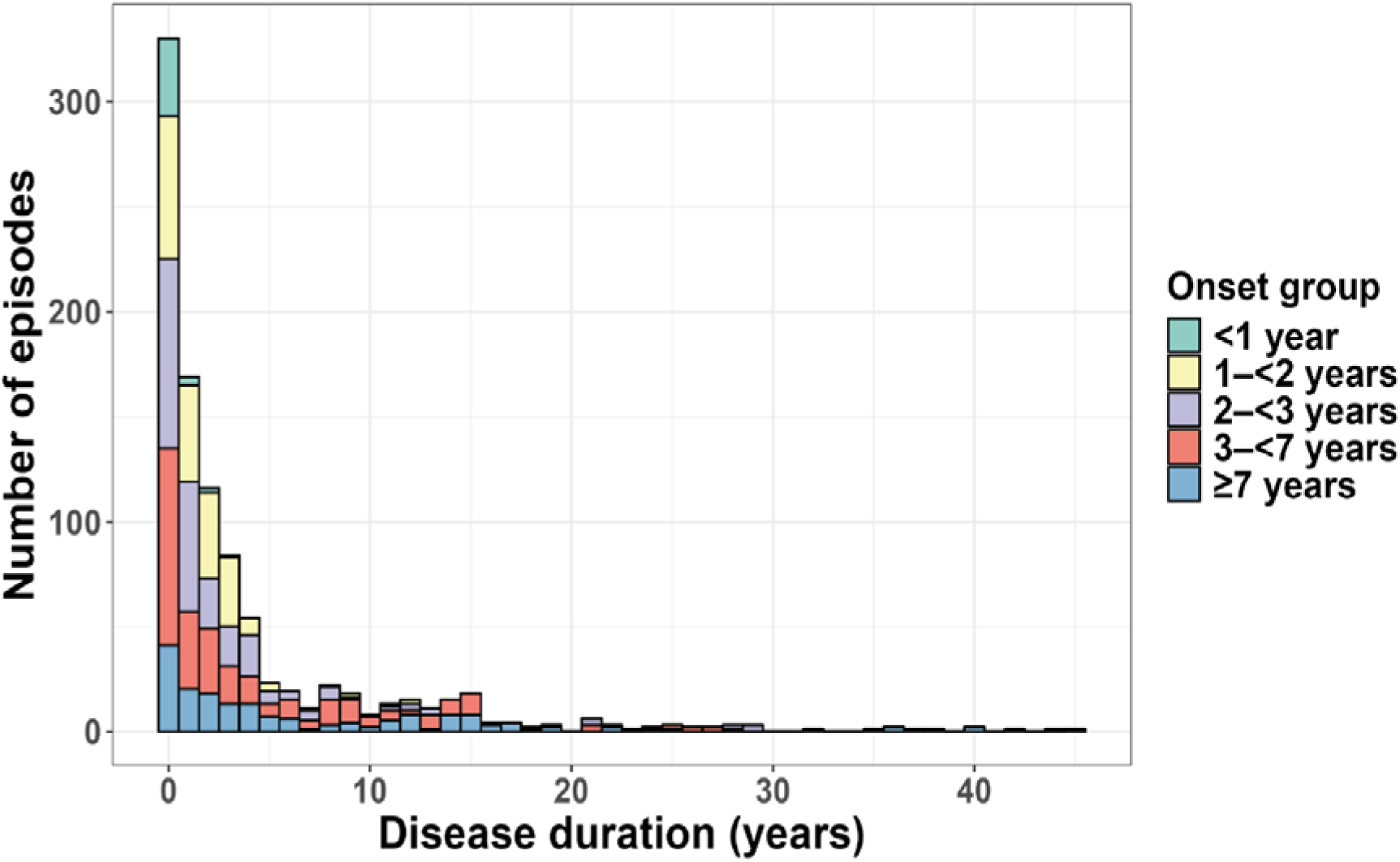
Number of episodes of rapid decline during the disease course by age of onset

### HUI multiscores

The longitudinal evolution of HUI multiscores was different across the age of onset categories, with patients with onset between 2 to <3 years having a faster disease progression than later onset patients (Figure 6). Of note, HUIs were not collected below 2 years of age. In all age of onset groups, patients showed highly diverse trajectories including rapid decline, improvement after rapid decline, periods of stability, or continued deterioration. Baseline imputation in the spaghetti plots helped visualize and assess disease progression, which is difficult in the analyses without imputation (Supplementary Figure 3). When summarized per ambulation status or death, HUI multiscores showed a distinct ordering, reflecting the association between multi-domain function and disease progression (Supplementary Table 1).

**Figure 6.**
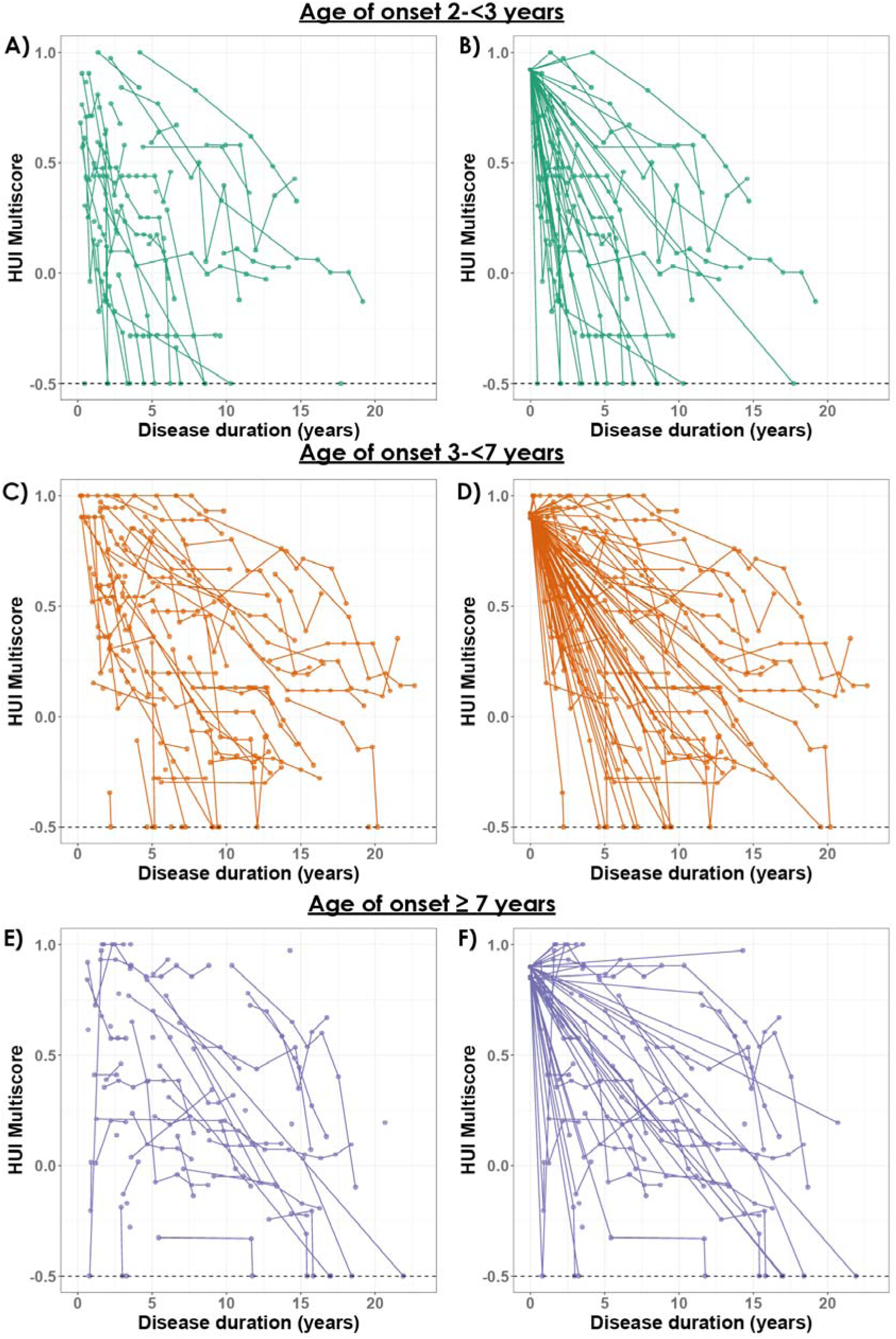
Panel HUI multiscores by the new age of onset categories The curves show HUI multiscores without baseline imputation for A) age of onset between 2–<3 years (199 HUIs of 41 patients), B) 3–<7 years (365 HUIs of 76 patients), and C) ≥7 years (203 HUIs of 59 patients) and HUI multiscores with baseline imputations for age of onset between D) 2–<3 years (158 HUIs of 41 patients), E) 3–<7 years (289 HUIs of 76 patients), and F) ≥7 years (159 HUIs of 59 patients).

## Discussion

VWM is an ultra-rare disease with a dismal prognosis. Multiple therapies are emerging^1^, but clinical trials are difficult to design and execute. Low patient numbers pose recruitment challenges and a placebo-controlled setup may be neither ethically acceptable nor feasible for patients and their families. The use of historical controls from a registry may represent a valuable alternative, but this approach also entails methodological challenges. The current study addresses these challenges, using the VWM registry as a case study to inform clinical trial design.

We first considered whether all patient types are suitable to serve as controls. One subpopulation comprises patients with comorbidities that may influence disease progression. They were excluded from the study, as they typically are from clinical trials. Asymptomatic or presymptomatic patients constitute another subpopulation. They cannot be used as historical controls as they have no known disease onset, a crucial item to determine disease duration and for stratification (see below). A third subpopulation comprises patients who experienced a single acute episode, often precipitated by relatively severe physical stressors, after which they returned to being free of neurological signs, suggesting that their “meant to be” age of onset is later. These patients might not be suitable controls as their age of onset in this initial episode would be earlier than the “meant to be” age of onset. This finding highlights the interplay between genetic susceptibility and environmental triggers underlying stress sensitivity in VWM: if the stressor is severe, it may provoke acute decline in milder cases with later “natural” disease onset. By contrast, some patients experience disease onset with a first episode of neurological decline, and progress directly from presymptomatic state to death. This is a common presentation among patients with early-infantile disease onset, but may also occur with later onset. This presentation is particularly unusual in adolescent or adult patients.^2^ Similar cases have been reported before.^27–29^ In hindsight, transient neurological signs and symptoms attributable to VWM may have been present, but this applies to all patients. We included these patients in the current study, but it is debatable whether such exceptional cases are representative of the natural disease course or should be excluded.

A second important question is whether the disease course has changed over time, whether historical data from all periods are useable, and if not, which period should be used. We assumed that general improvements in healthcare, such as better nutrition^30^, and faster accurate diagnosis after publication of the associated genes in 2001 and 2002^21,26^, followed by the implementation of preventive measures to avoid physical stressors, as well as increased awareness of the disease, may all have resulted in a milder disease course. We did observe temporal changes in disease course but, unexpectedly, the oldest historical controls, with onset before 1991, exhibited a milder disease than patients with more recent onset. This finding is likely attributable to selection bias: for patients with onset before 1991, only those who survived long enough to receive a DNA-based diagnosis could be included in the registry, leading to underrepresentation of rapidly progressive cases who had no DNA-confirmed diagnosis and did not enter the registry. This reflects another inherent limitation of registry-based recruitment: patients who die or are lost to follow-up before diagnosis or registry enrollment, are not captured. Therefore, patients with the poorest prognosis are less likely to be captured in a registry and patients with slower disease progression are overrepresented, skewing measures of the disease progression towards those of milder, slower-progressing cases.^31,32^

We did not find statistical evidence for drift in patients from 1991 onwards. The absence of statistical evidence is, however, limited by the low sample size. A combined argument with clinical reasoning stands stronger than statistical analysis alone. As the genes associated with VWM were published in 2002/2002, allowing a DNA-based diagnosis, and the usual care for VWM patients changed at that time, we would preferably use information from registry participants from 2002 onwards. If this yields insufficient patients to do any feasible analysis, the threshold can be changed to before 2002, although this might be at the cost of bias. Importantly, prioritizing more recent registry patients is in line with FDA and EMA guidelines.^33,34^

The third issue concerns selection of appropriate endpoints. Episodes of acute decline negatively affect the disease course^2^ and may appear an easily measurable trial endpoint. We found that the majority of patients experienced at least one episode of rapid decline during their disease course, but mostly at presentation. The low number of acute episodes after disease onset limits their suitability as endpoints for clinical trials, which typically have a duration of 1-2 years. Additionally, data validity may be suboptimal. Some families reported unlikely high numbers of episodes. Duplicate reporting may explain part of the problem, but also lack of formal definition of an episode of rapid decline makes it difficult for families to interpret consistently, potentially leading to over- and underreporting. So, episodes of rapid decline, as currently captured in the VWM registry, do not represent a reliable outcome measure. Ambulation and overall survival may represent more suitable endpoints. Our multi-state model analysis demonstrated that age of onset has a stronger association with ambulation with support than with survival. We believe that this is because patients generally attempt to maintain their walking ability for as long as possible, making loss of ambulation a well-defined and objective event. By contrast, survival is influenced by social, cultural, and healthcare-related factors that affect decisions regarding end of life, as patients can be kept alive through technical means for a long time. This limits the value of overall survival as an endpoint considerably, especially when participants from a trial are compared to historical cohorts.^35,36^ In line with this, we observed some patients with early-infantile onset who survived for extended periods despite severe or end-stage neurological impairment, managed by full supportive care.

Repeated measurements over time may be considered as endpoints. A major strength of the registry is the availability of longitudinal measurements of the HUI multiscore for numerous VWM patients over 20 years. These data demonstrate heterogeneous disease trajectories between age of onset groups and between patients within the same group. Patients may experience periods of stability, decline, and occasionally improvement. Consequently, improvements in clinical outcomes should be interpreted with caution in the absence of control data. A limitation of using longitudinal clinical scores is that patients are not assessed just before disease onset; the first assessment occurs after an established VWM diagnosis and registry enrollment. The first HUI measurement is typically several months after initial presentation, sometimes several years^2^. Consequently, as in many registries^32,37,38^, the VWM registry fails to capture the early disease phase. To address this issue and visualize what are the expected trajectories over time, we imputed reference HUI multiscores as baseline just prior to disease onset, as previously^2^, and demonstrated that this approach is not only logical but also allows the modeling of the theoretical disease trajectories from clinical presentation on.

The fourth consideration concerns patient stratification. Age of onset has been shown to be the best predictor of disease severity^2^ and stratification for age of onset is important for trials^7^. The previous age of onset categories were primarily defined on clinical judgement.^2^ The integration of data-driven analyses with clinical expertise enabled identification of new age of onset categories better capturing phenotypic differences across the categories. However, given the low incidence of VWM and a limited study sample size, stratification using the previous or newly defined age of onset categories may not be feasible. Therefore, stratification into two categories, with onset before 3 years and at 3 years or older, may represent a pragmatic approach, distinguishing more severe from milder disease. Stratification for sex should also be considered. Since long, overrepresentation of females has been observed among adult patients with VWM.^2,39,40^ In a previous natural history study^2^, sex-related differences were not confirmed statistically. We reopened the question and identified an effect of sex on disease severity. This effect is expressed in difference in age of onset between males and females, with females presenting at later age and exhibiting milder disease than males. After adjusting for age of onset, the remaining effect of sex on disease progression was decreased and no longer statistically significant. For clinical trials, it seems that stratification for sex is not necessary after stratification for age of onset, but sensitivity analyses accounting for sex as a potential confounder are advisable.

In conclusion, the current study shows that longitudinal data from a large cohort of untreated patients can provide crucial information for trial design and analysis in ultra-rare diseases. The study highlights key considerations for the use of historical controls and underscores the need for collaborative efforts to establish registries for ultra-rare diseases before therapies become available.

## Declarations

## Acknowledgements

MSvdK is a members of the European Reference Network for Rare Neurological Disorders (ERN-RND), project ID 739510. The authors are grateful to the contributions of all physicians and specialists who provided clinical data on their patients.

## Author contributions

RJvV and EG conducted the data analyses and drafted the manuscript. MSvDK and RvE supervised the project. All authors contributed to the review and revision of the manuscript.

## Data availability statement

The data that support the findings of this study are available on request from the corresponding author. The data are not publicly available due to privacy or ethical restrictions.

## Disclosures

RJvV, EG, EMCH, MDS, MCP, JB, RvE report no disclosures relevant to the manuscript.

MSvdK: co-investigator for Calico (VWM, Fosigotifator trial) and co-investigator for Ionis (Alexander disease trial). She is on patents EP003618817B1, P112686US00 and P112686CA00 on “therapeutic effects of Guanabenz treatment in vanishing white matter” for the VU University Medical Center, Amsterdam, The Netherlands. She is the initiator and principal investigator of the Guanabenz trial (https://www.clinicaltrialsregister.eu/ctr-search/trial/2017-001438-25/NL), with permission of the Dutch national ethics committee (CCMO, NL61627.000.18).

## Funding

NWO, Spinoza premie

## Notes

### Author Declarations

Dutch national ethics committee (CCMO, NL61627.000.18).

