## Supplementary Materials for "The vanishing white matter registry: a case study of historical controls in clinical trial design"

**Supplementary Table 1. HUI multiscore score distribution summarized per ambulation status**

| **Ambulation status¹** | **n²** | **Mean (±SD)**  **HUI multiscore** | **Median (Q1, Q3) HUI multiscore** |
| --- | --- | --- | --- |
| **Independent ambulation** | 229 | 0.64 (±0.31) | 0.67 (0.44, 0.91) |
| **Ambulation without support lost** | 167 | 0.43 (±0.26) | 0.44 (0.20, 0.59) |
| **Ambulation with support lost** | 212 | 0.07 (±0.26) | 0.05 (-0.02, 0.34) |
| **Death** | 66 | -0.5^3^ | -0.5^3^ |

¹Imputed reference scores were not included in this analysis.²Note that patients can transition through multiple ambulation states, starting with independent ambulation and ending at death. ^3^Death was assigned a fixed score of −0.5, with no variability. SD= standard deviation.

**Supplementary Figure 1**. **Kaplan–Meier curves with year of onset and loss of ambulation without support**

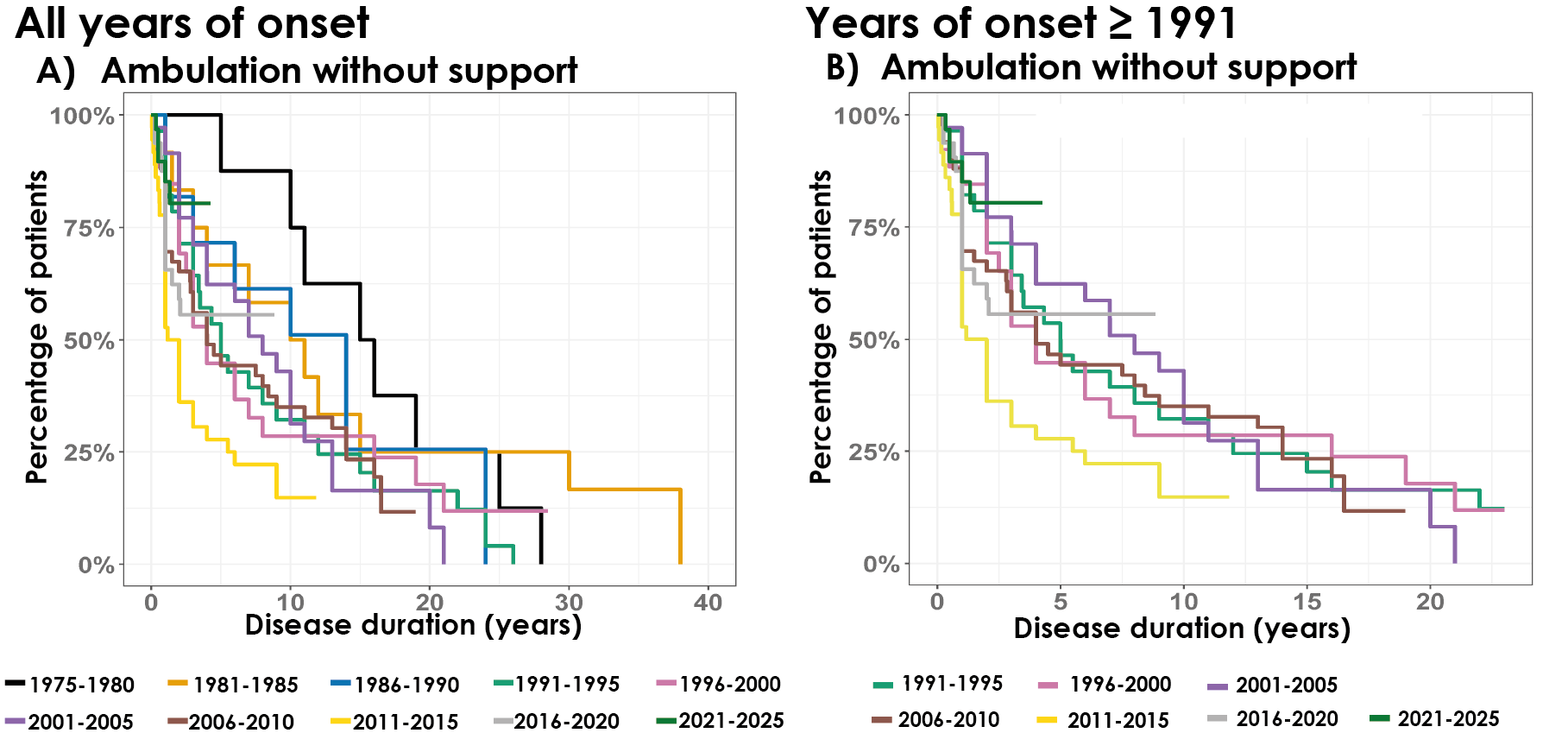

A) Overall time to loss of ambulation without support by year of onset: 1975-1980 (n=8); 1981-1985 (n=12), 1986–1990 (n=11), 1991–1995 (n=28), 1996–2000 (n=26), 2001-2005 (n=36), 2006–2010 (n =50), 2011–2015 (n=36), 2016–2020 (n=32), 2021–2025 (n=32); **B**) Time to loss of ambulation without support in patients with onset from 1991 onwards: 1991-1995 (n=28), 1996-2000 (n=26), 2001-2005 (n=36), 2006–2010 (n=50), 2011–2015 (n=36), 2016–2020 (n=32), 2020–2025 (n=32).

**Supplementary Figure 2. Predictions of the multi-state model with disease transitions**: independent ambulation, ambulation with support lost, and death. Transition probabilities from one state to another.
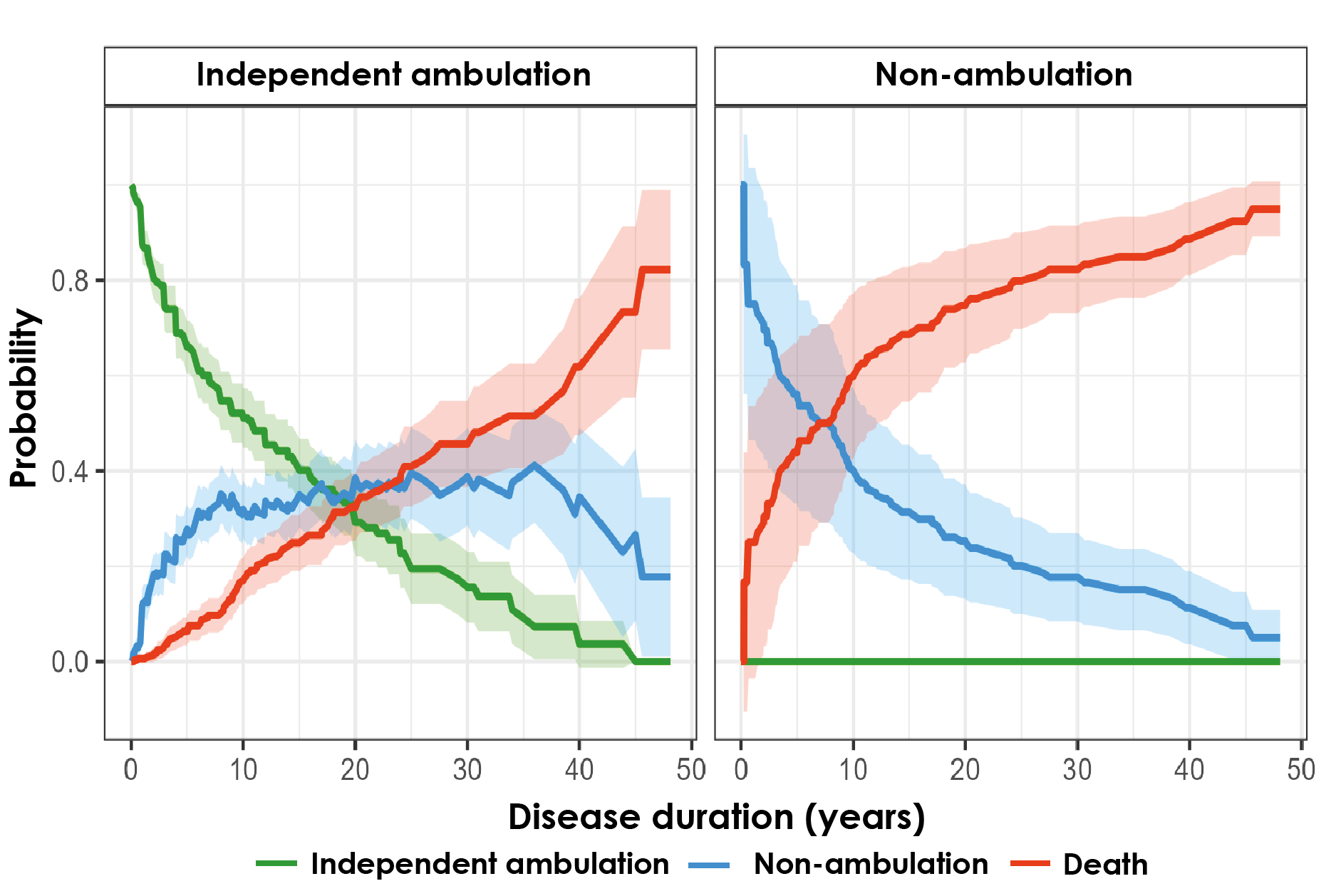

**Supplementary Figure 3. Predictions of HUI multiscores over time by age of onset**

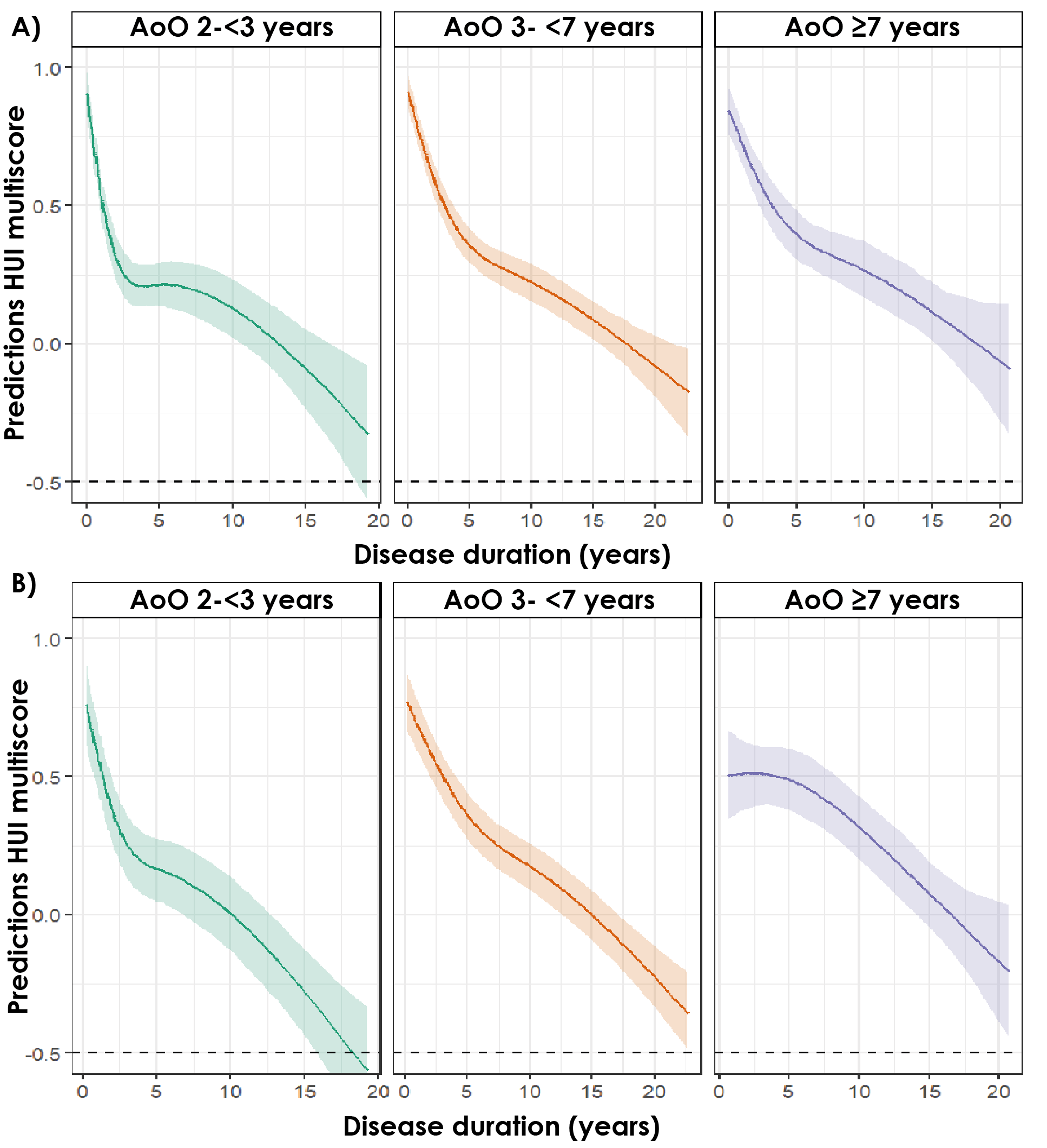

These curves represent the estimated average trajectories for a patient within each age-of-onset group. Panel  **A**) shows HUI multiscore with baseline imputation and panel **B**) shows HUI multiscores without baseline imputation. In both models, HUI multiscores were modelled with natural splines.

AoO, age of onset.
